# Transmission dynamics and epidemiological characteristics of Delta variant infections in China

**DOI:** 10.1101/2021.08.12.21261991

**Authors:** Min Kang, Hualei Xin, Jun Yuan, Sheikh Taslim Ali, Zimian Liang, Jiayi Zhang, Ting Hu, Eric H. Y. Lau, Yingtao Zhang, Meng Zhang, Benjamin J. Cowling, Yan Li, Peng Wu

## Abstract

**Background:** The Delta variant of SARS-CoV-2 has become predominant globally. We evaluated the transmission dynamics and epidemiological characteristics of the Delta variant in an outbreak in southern China.

**Methods:** Data on confirmed cases and their close contacts were retrospectively collected from the outbreak that occurred in Guangdong, China in May-June 2021. Key epidemiological parameters, temporal trend of viral loads and secondary attack rates were estimated and compared between the Delta variant and the wild-type SARS-CoV-2 virus. We also evaluated the association of vaccination with viral load and transmission.

**Results:** We identified 167 patients infected with the Delta variant in the Guangdong outbreak. The mean estimates of the latent period and the incubation period were 4.0 days and 5.8 days, respectively. A relatively higher viral load was observed in Delta cases than in wild-type infections. The secondary attack rate among close contacts of Delta cases was 1.4%, and 73.9% (95% confidence interval: 67.2%, 81.3%) of the transmissions occurred before onset. Index cases without vaccination (OR: 2.84, 95% confidence interval: 1.19, 8.45) or with one dose of vaccination (OR: 6.02, 95% confidence interval: 2.45, 18.16) were more likely to transmit infection to their contacts than those who had received 2 doses of vaccination.

**Discussion:** Patients infected with the Delta variant had more rapid symptom onset. The shorter and time-varying serial interval should be accounted in estimation of reproductive numbers. The higher viral load and higher risk of pre-symptomatic transmission indicated the challenges in control of infections with the Delta variant.

## INTRODUCTION

The SARS-CoV-2 Pango lineage B.1.617.2, also known as the Delta variant, is a variant of SARS-CoV-2 first detected in India on 7 September 2020 (1). It was classified by the World Health Organization as a “Variant of Concern” on 11 May 2021, and has been rapidly outcompeting other variants of SARS-CoV-2 and becoming predominant in many locations around the world. As of 3 August 2021, a total of 135 countries have reported cases of the Delta variant, and over 80% of new infections globally were expected to be due to Delta since mid-June (1, 2).

Compared to the wild-type virus, the Delta variant has 9-10 characteristic mutations including T19R, G142D, 156del, 157del, R158G, L452R, T478K, D614G, P681R, and D950N which could be responsible for competitive advantages against other variants (3). Residue 452 spike mutation located at the receptor binding domain may increase capability of immune evasion and resistance to antibody neutralization, and P681R in the S1/S2 regions of S gene could influence proteolytic processing (4). All these mutations could result in increased affinity of ACE2 and resistance to antibody neutralization therefore leading to increases in transmissibility (4). The basic reproduction number (R_0_) of Delta variant was suggested to be 55%-97% higher than other variants (2).

On 21 May 2021 the first local Delta case in mainland China was identified in Guangdong province. A local outbreak occurred in the following days and weeks, and the gene sequence analysis showed that all cases identified in this outbreak were infected with the Delta variant and could be traced back to the index case (5). Aggressive case finding strategy including multiple comprehensive large-scale nucleic acid tests in high-risk communities, routine PCR testing for close contacts quarantined in designated places and nucleic acid screening among inpatients and outpatients in clinical institutions, had been strictly implemented aiming to identify all infected persons and rapidly control this outbreak. This provides uniquely rich epidemiological data on infections with the Delta variant. In this study, we aimed to explore the transmission dynamics and epidemiological characteristics of the Delta variant outbreak in China. As the coverage of COVID-19 vaccination has increased substantially in China since March 2021, we were able to examine the associations between vaccination and virus shedding and transmission.

## METHODS

### Data collection

We retrospectively collected information on all laboratory-confirmed symptomatic and asymptomatic cases with Delta (B.1.617.2) variant infection from the outbreak in Guangdong province in May and June 2021. To estimate the latent period distribution, we collected individual information on the first and last dates of exposure (exposure window), and the repeated laboratory testing dates of last negative PCR test (lower bound of viral shedding) and first positive PCR test (upper bound of viral shedding) which provide a window during which detectable virus shedding began. We also obtained illness onset dates for incubation period estimation. We reconstructed the transmission pairs from available illness onset dates for both infectors and infectees to estimate the serial interval distribution, the infectiousness profile, and the proportion of transmission occurring prior to symptom onset. Severity status including asymptomatic, mild, moderate, severe and critical were collected for each case, along with other information such as sex, age, pre-existing underlying conditions, vaccination status, and exposure duration. We also collected information on close contacts of the confirmed Delta cases to estimate secondary attack rates and identify predictors of infection.

Comprehensive large-scale nucleic acid-testing strategies, including community-wide PCR testing, routine test among concentrated quarantine close contacts and daily test for inpatients, were implemented in every local COVID-19 outbreak in China since April 2020 (6). For each case, serial samples were collected and tested for both N-gene and OR-gene from the date of first positive PCR test until discharge from hospital. To understand the temporal dynamics of viral RNA shedding for the Delta variant, we obtained serial cycle threshold (*Ct*) values for each case for N-gene with throat swabs from the first time of positive test (*Ct* value <40). To make a comparison of viral loads between the Delta variant and the wild-type variant, we used data of wild-type SARS-CoV-2 infections that were identified in Guangzhou, China in early 2020 from a published paper on individual cases with daily test results (7).

### Case definitions

A patient is confirmed as a COVID-19 case based on a positive result of PCR for SARS-CoV-2 with respiratory specimens. Virus strains in this study were determined by the sequenced genome and were classified based on the “Pango lineages” rule (8). The time interval between infection and becoming infectious is defined as the latent period, that could be compared with the incubation period which describes the time duration between infection and symptom onset. The latent period is typically proxied by the time from infection until an infected person has virus shedding that is detectable by PCR, and can be shorter than the incubation period for some COVID-19 cases when virus shedding becomes detectable prior to symptom onset. The serial interval, defined as the time interval between successive symptom onsets in a transmission chain, is an important parameter for estimating many other key epidemiological parameters, such as R_0_, the expected number of secondary cases generated from one primary case in a completely susceptible population, and the instantaneous reproduction number (R_t_) which describes the expected number of secondary cases caused by one typical primary case at time *t*. The infectiousness profile of COVID-19 describes the duration and intensity of infectiousness of infected cases which imply the probability of transmission during the infectious period.

We assessed the clinical severity of COVID-19 cases via clinical classification into asymptomatic, mild, moderate, severe and critical following the Guidelines in Diagnosis and Treatment of COVID-19 (8^th^ version) published by National Health Commission since 15 April 2021 (9).

Close contacts were defined as individuals who were exposed to symptomatic COVID-19 cases within two days before their illness onset, or exposed to asymptomatic cases at close proximity (<1 meter) without wearing proper personal protection equipment within two days before their sampling dates of the first positive samples for SARS-CoV-2. Close contacts were classified as household and extended family, social, community and healthcare contacts based on the definitions previous published by *Sun et al* (10). Cases were considered having effective 1-dose vaccination if the start date of exposure was 10 days after the first dose of vaccination or later, or having effective 2-dose vaccination if the start date of exposure was 14 days after the second dose of vaccination or later (11, 12).

### Statistical analysis

We used a maximum likelihood-based inferential method to estimate the distributions of latent period, incubation period and serial interval and the infectiousness profile of confirmed COVID-19 cases by fitting Gamma distributions. We accounted for the interval censoring of exposure and viral shedding windows when estimating the latent period and incubation period distributions. To estimate the infectiousness profile for symptomatic cases, we used a method previously published by *He et al*. (7) which considered the serial interval as a convolution between the infectiousness profile and the incubation period and allowed for an early occurrence of infectiousness before symptom onset.

The time-varying forward serial intervals (13, 14) and daily numbers of cases were used to estimate the daily R_t_ by applying the statistical methods developed by *Cori et al* (15). The serial interval distribution and R_0_ were obtained by using mean estimates of the serial interval and R_t_ during the exponential growth phase of the Delta outbreak (14).

The overall temporal trend of *Ct* values for N-gene for Delta cases was analyzed by day of illness onset. To aid visualization, smoothing splines using generalized additive models (GAMs) (including days of illness onset as the only predictor) were fitted to the *Ct* values to characterize the overall trend for the Delta variant. To make a comparison between Delta and wild-type, we also fitted the temporal trend of *Ct* values for the Delta variant and wild-type separately by excluding severe and critical and vaccinated Delta cases, because no severe or critical or vaccinated cases were identified in wild-type cases. To evaluate the impact of vaccination on viral loads among Delta cases, we fitted a multivariate GAMs by including variables of vaccination (1: without vaccination, 2: with one or two dose of vaccine), days of illness onset, age and disease severity. Temporal trend of predicted *Ct* values from the GAMs model was presented and compared using box plots for vaccinated and unvaccinated cases separately.

Close contacts of confirmed COVID-19 cases infected with Delta variant with a solely possible source of infection for each close contact were included for analyzing. The overall secondary attack rate was calculated by dividing the number of infections by the total number of close contacts. To assess the effectiveness of vaccination against transmission, a stepwise logistic regression model was fitted by including age, sex, disease severity of the index, COVID-19 vaccination for index cases, COVID-19 vaccination for close contacts, type of contact, presence of exposure on the symptom onsets of index cases and duration of exposure. Non-parametric and parametric bootstrap approach with 1000 resamples was used to assess the uncertainty of each estimated parameter. Analyses were carried out using R version 4.0.3 (R Foundation for Statistical Computing, Vienna, Austria).

## RESULTS

As of 18 June 2021, 167 Delta cases were identified in the outbreak in Guangdong. Sixty-nine (41.3%) were male. The median age was 47.0 years (interquartile range [IQR]: 31.0, 66.5) with 22 (13.2%) cases aged under 15 years and 44 (26.3%) over 65 years. The number of asymptomatic, mild, normal and severe or critical was 8 (4.8%), 29 (17.4%), 111 (66.5%) and 19 (11.4%), respectively, with no reported deaths. Sixteen (9.6%) cases received 2 doses of the inactivated COVID-19 vaccine and 30 cases (18.0%) received one vaccine dose.

We examined data from 101 confirmed Delta cases with sufficient information to estimate the time window for infection and the time window for the start of viral shedding. The mean latent period was estimated to be 4.0 days (95% confidence interval [CI]: 3.5, 4.4). Ninety-five percent of the Delta cases started shedding virus within 8.2 days (95% CI: 7.1, 9.3) after infection (Figure 1A). The mean incubation period estimated from 95 symptomatic Delta cases was 5.8 days (95% CI: 5.2, 6.4). The 95^th^ percentile of the incubation period for Delta was 11.5 days (95% CI: 10.1, 13.0) (Figure 1B).

**Figure 1.**
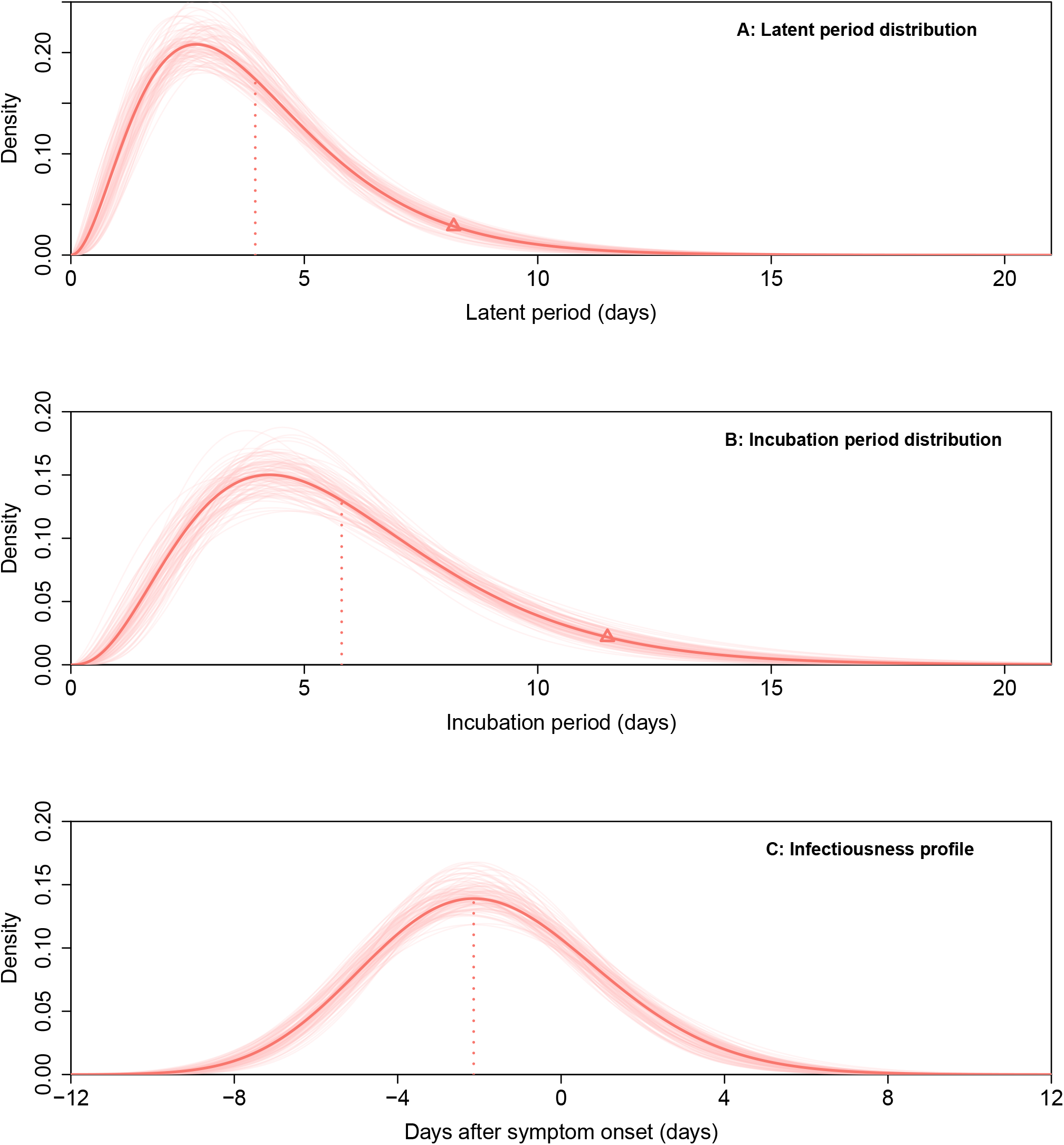
The key epidemiologic time-delay distributions of Delta variant (red lines). (A) The latent period distribution. (B) The incubation period distribution. (C) The infectiousness distribution. Vertical dotted lines show the mean estimates for latent period distribution and incubation period distribution, and the peak estimate for infectiousness profile. The triangles in A and B indicate the 95^th^ percentiles for respective parameters.

We used data from 94 transmission pairs of Delta cases with a reported onset date to estimate the infectiousness profile by allowing for transmission before symptom onset. We estimated that 2.7% (95% CI: 1.0%, 5.0%) of transmission occurred prior to 7 days before illness onset, 22.5% (95% CI: 16.0%, 30.0%) started to become infectious 4 days before illness onset, and the infectiousness peaked at 2.1 days (95% CI: 1.5, 2.7) before onset and then dropped gradually, with 73.9% (95% CI: 67.2%, 81.3%) of transmission occurred before illness onset and 97.1% (95% CI: 94.4%, 99.0%) of transmission occurred within 4 days after illness onset (Figure 1C).

The estimated forward serial intervals decreased from 6.1 days (95% CI: 5.2, 7.1) on 25 May 2021 to 4.0 days (95% CI: 3.1, 5.0) on 18 June 2021 in the Delta outbreak (Figure 2B). By using the time varying forward serial intervals and case incidence data, we estimated that the R_t_ dropped rapidly from 9.3 (95% CI: 7.7, 11.6) on 25 May 2021 to 0.48 (95% CI: 0.42, 0.57) on 18 June 2021, and had been below 1 since 9 June 2021 (Figure 2C). During the same time period, the estimated infectiousness peak shifted from 0.23 days (95% CI: 0.20, 0.26) after illness onset to 2.14 days (95% CI: 1.52, 2.70) before illness onset based on the time varying serial interval (Figure 2D). By using the daily estimates of forward serial interval and R_t_ during the exponential growth phase before 27 May 2021, the initial forward serial interval was estimated to be 5.8 days (95% CI: 5.2, 6.1) (Figure 2B), and the R_0_ was 6.4 (95% CI: 3.7, 9.3) (Figure 2C).

**Figure 2.**
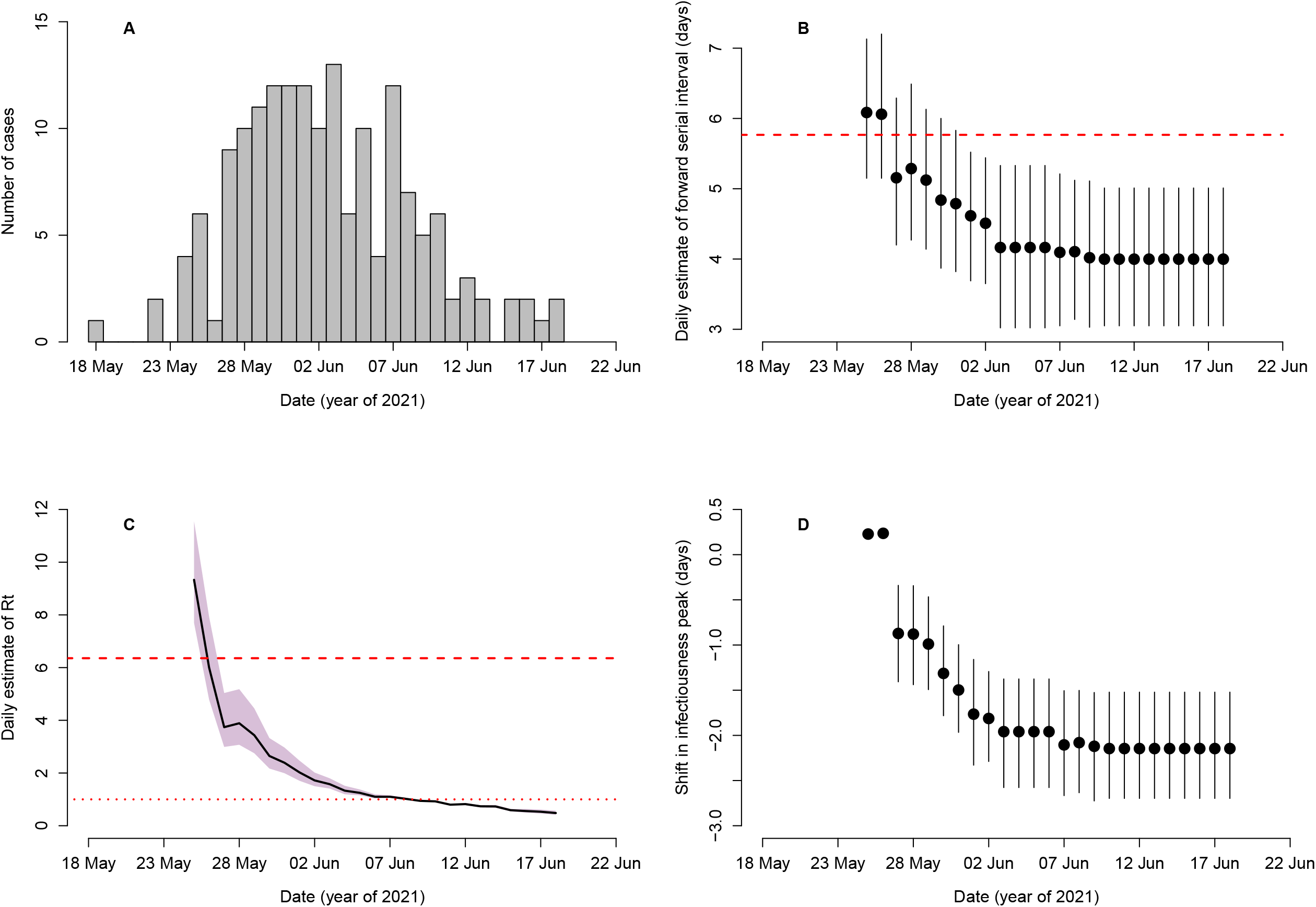
The daily estimates of forward serial interval, instantaneous reproduction number (R_t_) and infectiousness peak. (A) The epidemic curve based on symptom onset dates of all cases in the Delta outbreak in Guangdong province. (B) The forward estimates of the serial interval over time. The dots represent the daily mean estimates of serial interval, and the vertical line segments represent the 95% CIs. The dashed line represents the initial (from transmission pairs with illness onsets of infectors before May 27, 2021) forward serial interval (5.8 days) (C) Mean estimates of R_t_ (line) and the 95% CIs (shaded area) based on the daily forward estimates of serial interval. The dashed line represents the basic reproduction number R_0_ (6.4). The dotted line indicates R_t_=1. (D) Estimates of the daily infectiousness peak after illness onset, based on daily estimates of the serial interval. The dots represent the daily mean estimates of infectiousness peak, and the vertical line segments represent the 95% CIs.

In total, 1314 throat swabs collected between 4 days before and 34 days after illness onset were tested for 159 Delta cases. High viral loads were maintained between 4 days before onset and 7 days after onset, then decreased gradually to a low but detectable level until about Day 20 (Figure 3A). To compare viral loads between Delta and wild-type, we identified 94 cases with the median age of 46 years (IQR: 33, 61) infected with the wild-type virus in Guangzhou city, Guangdong province in China between 21 January 2020 and 14 February 2020. Among those, 47 (50.0%) were male, the number of asymptomatic, mild and normal were 2 (2.1%), 30 (31.9%) and 61 (64.9%), respectively, and no severe or critical cases were identified. None of these cases received COVID-19 vaccination. In total 406 throat swab samples were collected and tested on the illness onset day and 31 days after onset for the 94 wild-type cases. After excluding severe and critical cases and vaccinated cases, we found during the period with a high viral load (0 to 7 days after onset), the median *Ct* values were 23.0 (IQR: 19.3-28.6) for N gene of the Delta variant, significantly lower than the values of the wild-type N gene (median: 36.5, IQR: 33.0-40.0) (Figures 3B). Results of the GAMs revealed that the *Ct* values of Delta cases who had one dose or two doses of vaccination were on average 0.97 (95% CI: 0.19, 1.76) higher than unvaccinated cases after adjusting for days of illness onset, age and disease severity (Figures 3C).

**Figure 3.**
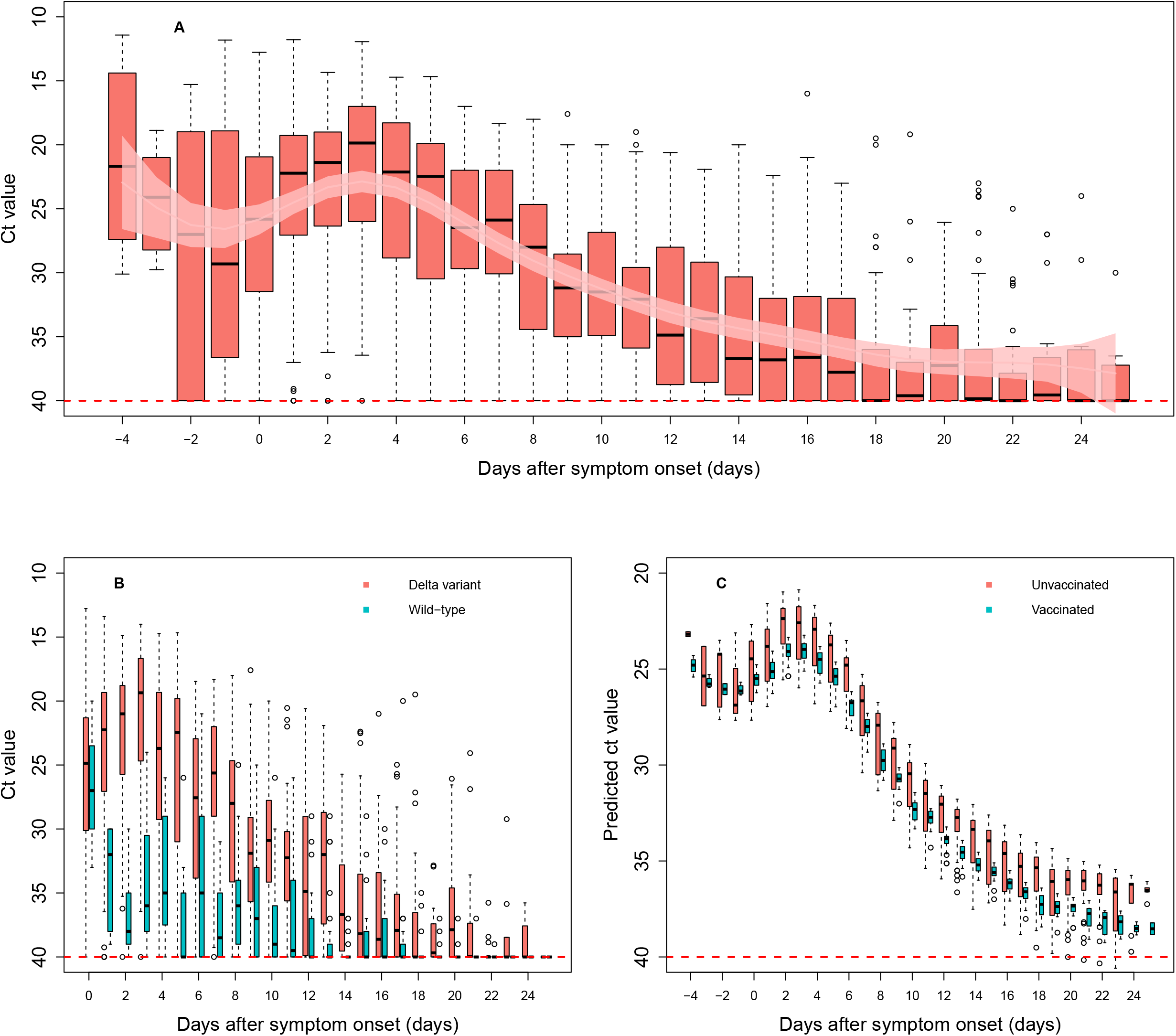
Temporal patterns of viral shedding for the Delta variant and the wild-type SARS-CoV-2 virus. (A) Boxplot of threshold cycle (*Ct*) for N gene for all cases infected with the Delta variant. Light red curve and light pink shaded area indicate the fitted Ct values and the 95% CIs estimated from the generalized additive models (GAMs). (B) Boxplot of *Ct* values for N gene of Delta (red) and wide-type (blue) against time of sample collection relative to the date of illness onset. The data for the Delta variant excluded severe, critical and vaccinated cases. (C) Boxplot of the predicted *Ct* values from the multivariate GAMs for unvaccinated (red) and vaccinated (blue, with one or two doses) cases.

To evaluate individual infection risk and the effectiveness of vaccination on transmission for the Delta variant, we analyzed infections among 5153 individuals who were close contacts of 73 COVID-19 cases. The overall secondary attack rate was 1.4% (95% CI: 1.1%, 1.8%) in the contacts. The stepwise regression model showed that a high infection risk was among those in older age (OR: 1.02, 95% CI: 1.01, 1.03), exposed to an index case without vaccination (OR: 2.84, 95% CI: 1.19, 8.45) or with 1 dose of vaccination (OR: 6.02, 95% CI: 2.45, 18.16), and being household and extended family contacts (OR: 40, 95% CI: 24, 66) (Table 1).

**Table 1.** Secondary attack rate for close contacts of the Delta infectors, and the risk factors associated with the occurrence of infection based on backward logistic regression.

| Characteristics | No. of close contacts | No. of infections (%) | Adjusted OR (95% CI) |
| --- | --- | --- | --- |
| **Overall** | 5153 | 73 (1.4) |  |
| **Sex** |  |  |  |
| Male | 2553 | 29 (1.1) |  |
| Female | 2600 | 44 (1.7) |  |
| **Age, years, median (IQR <sup>a</sup>)** | 47.0 (31, 66.5) |  | 1.02 (1.01-1.03) |
| 0- | 424 | 12 (2.8) |  |
| 15- | 2683 | 18 (0.7) |  |
| 45- | 1521 | 27 (1.8) |  |
| 65- | 525 | 16 (3.0) |  |
| **Type of index cases** |  |  |  |
| Asymptomatic and mild | 1809 | 12 (0.7) |  |
| Normal, Severe or critical | 3344 | 61 (1.8) |  |
| **COVID-19 Vaccine dose of index cases** |  |  |  |
| 0 | 2892 | 37 (1.3) | 2.84 (1.19-8.45) |
| 1<sup>b</sup> | 1110 | 31 (2.8) | 6.02 (2.45-18.16) |
| 2<sup>c</sup> | 1151 | 5 (0.4) | Referent |
| **COVID-19 Vaccine dose of contacts** |  |  |  |
| 0 | 2844 | 48 (1.7) |  |
| 1<sup>b</sup> | 1459 | 17 (1.2) |  |
| 2<sup>c</sup> | 850 | 8 (0.9) |  |
| **Type of contact** |  |  |  |
| Household and extended family | 173 | 38 (22.0) | 40 (24-66) |
| Others | 4980 | 35 (0.7) | Referent |
| **Exposure to an index case at onset <sup>d</sup>** |  |  |  |
| Yes | 2106 | 49 (2.3) |  |
| No | 3047 | 24 (0.8) |  |
| **Duration of exposure, days, mean (sd)** | 7.8 (3.8) |  |  |
| 1- | 1092 | 7 (0.6) |  |
| 6- | 4061 | 66 (1.6) |  |

<sup>a</sup> IQR: interquartile range
<sup>b</sup> first COVID-19 vaccine dose 10 days before the first day of possible exposure to an infector
<sup>c</sup> second COVID-19 vaccine dose was given 14 days before the first day of possible exposure to an infector
<sup>d</sup> close contacts were exposed to an index at the time of the onset day of the index

## DISCUSSION

Our study provided a comprehensive assessment of the epidemiological characteristics of the Delta variant. Higher transmissibility was demonstrated for the Delta variant, as indicated by a higher reproduction number, shorter latent and incubation periods, and shorter serial intervals compared to the wild-type SARS-CoV-2 (5, 13, 16-18). We observed higher viral loads in cases infected with the Delta variant which might contribute to more rapid and intense transmission. In addition, we found the inactivated vaccines could effectively reduce viral loads in cases infected with the Delta variant and further lead to lower transmissibility.

We estimated the time varying forward serial intervals which considered the temporal dynamics of the disease transmission in an outbreak (13, 14). The R_0_ estimated for the Delta variant was 6.4 which was substantially higher than the R_0_ of the wild-type virus at the start of the pandemic (16, 19). Estimation of the reproduction number could be underestimated due to unobserved infections and neglecting the changes in the forward serial interval distribution during the period of epidemic (14). In the Delta outbreak in China, active and aggressive case-finding strategies using multiple PCR tests were implemented, which was able to identify most infected persons including asymptomatic cases. With the shorter latent and incubation period, and higher secondary attack rate among household and extended family contacts, we believe multiple and more stringent interventions are needed to control epidemics of the Delta variant. During the Delta outbreak in China, the local government had implemented individual-based interventions such as case isolation, contact tracing and quarantine, as well as population-level physical distancing measures such as lockdowns and confinement (10, 13). More importantly, various community-wide PCR testing and routine testing programmes among quarantined close contacts were aligned with the measures of contact tracing and lockdown, aiming to identify and isolate the cases as early as possible and interrupt transmission chains. The rapid drop in R_t_ within a week (Figure 2C) indicated the effectiveness of these interventions.

We estimated that the 73.9% of transmissions occurred pre-symptomatically for the Delta variant, which was higher than other variants (7, 20, 21), suggesting a higher transmission potential of Delta cases before detection which was further supported by the high viral loads at least 4 days before illness onset shown in our study. The high risk of transmission particularly before onset indicated the need to expand contact tracing to a wider group of contacts and perhaps to a longer time scale in order to control the epidemic caused by the Delta variant (7, 22). However, for areas with a high prevalence of COVID-19, complete contact tracing and quarantine outside the home may be infeasible as the number of contacts is always several folds the number of infections (10). Physical distancing such as self-isolation and home quarantine is more suitable in these areas. However, society-wide physical distancing measures might increase transmission risk at household settings (10, 20). Our study showed that the secondary attack rate (22.0%) among household close contacts of Delta cases was higher than the rate obtained in 2020 (12.4%) in the same location with wild-type infections (23).

We found that the viral load was higher in cases of the Delta variant than cases of the wild-type virus, indicating a potentially higher infection rate per contact for the Delta (24). In addition, patients infected with the Delta variant maintained a high viral load from 4 days before illness onset. Besides, compared to the wild type, patients infected with Delta had a slower decline in viral load towards the detection threshold of the PCR test (Figure 3B), likely leading to a longer infectious period (24). Escape of the Delta variant from immunity induced by wild-type variants (25) suggests that the herd immunity threshold needed to suppress transmission of the wild-type virus may not be sufficient to control spread of the Delta variant (24). Higher burden of SARS-CoV-2 is expected in the future given the increasing predominance of the Delta variant all over the world. Additional booster doses of vaccination might be able to increase protection against the Delta variant.

The effectiveness of the current vaccines from Pfizer-BioNTech and Oxford-AstraZeneca appeared to diminish against infections with the Delta variant (25, 26). However, the efficacy of the vaccines against transmission, which is another important indicator of their impact (27), has rarely been reported. In this study, we observed that the *Ct* values among Delta cases with one or two doses of vaccination were on average 0.97 higher than the unvaccinated cases, indicating approximately 3-fold decrease in the quantity of viral RNA copies (28). The vaccinated Delta cases in our study with a decreased viral load might have a reduced transmission potential given that viral RNA load of SAS-CoV-2 was independently associated with the shedding of transmissible viruses (29). The effectiveness of inactivated vaccines against transmission of the Delta variant demonstrated the importance of increasing vaccination coverage in mitigating COVID-19 (30).

Our study had several limitations. Self-reported symptom onset might bias estimates of the parameters, e.g., leading to an overestimation of the incubation period if patients tended to remember the later days with symptoms. Second, the *Ct* values used in our study were obtained from different diagnostic kits which shared the same detection threshold but perhaps with different sensitivity and/or specificity. Finally, in estimation of the serial interval, transmission pairs with asymptomatic cases would be excluded due to absence of symptom onset dates, which however might have biased the estimates of the reproduction number by neglecting the impact of asymptomatic transmission.

In conclusion, the Delta variant demonstrated a higher transmissibility compared to the wild type of SARS-CoV-2. An extension of contact tracing period to perhaps four days prior to symptom onset may be needed considering the high proportion of pre-symptomatic transmission and the high viral load before onset in infections with the Delta variant. Inactivated vaccines appeared to be effective in reducing transmission of Delta infections and a high vaccination coverage should be pursued to reduce the burden of COVID-19 pandemic.

## Data Availability

Due to privacy and ethical concerns, the raw data cannot be made available.

## ACKNOWLEDGMENTS

The authors thank Julie Au for technical support.

